# Multireader Evaluation of Radiologist Performance for COVID-19 Detection on Emergency Department Chest Radiographs

**DOI:** 10.1101/2021.10.20.21265278

**Authors:** Judy W. Gichoya, Priyanshu Sinha, Melissa Davis, Jeffrey W. Dunkle, Scott A. Hamlin, Keith D. Herr, Carrie N. Hoff, Haley P. Letter, Christopher R. McAdams, Gregory D. Puthoff, Kevin L. Smith, Scott D. Steenburg, Imon Banerjee, Hari Trivedi

## Abstract

**Background:** Chest radiographs (CXR) are frequently used as a screening tool for patients with suspected COVID-19 infection pending reverse transcriptase polymerase chain reaction (RT-PCR) results, despite recommendations against this. We evaluated radiologist performance for COVID-19 diagnosis on CXR at the time of patient presentation in the Emergency Department (ED).

**Materials and Methods:** We extracted RT-PCR results, clinical history, and CXRs of all patients from a single institution between March and June 2020. 984 RT-PCR positive and 1043 RT-PCR negative radiographs were reviewed by 10 emergency radiologists from 4 academic centers. 100 cases were read by all radiologists and 1927 cases by 2 radiologists. Each radiologist chose the single best label per case: *Normal, COVID-19, Other – Infectious, Other – Noninfectious, Non-diagnostic*, and *Endotracheal Tube*. Cases labeled with *endotracheal tube* or *non-diagnostic* were excluded. Remaining cases were analyzed for label distribution, clinical history, and inter-reader agreement.

**Results:** 1727 radiographs (732 RT-PCR positive, 995 RT-PCR negative) were included from 1,594 patients (51.2% male, 48.8% female, age 59 ± 19 years). For 89 cases read by all readers, there was poor agreement for RT-PCR positive (Fleiss Score 0.36) and negative (Fleiss Score 0.46) exams. Agreement between two readers on 1,638 cases was 54.2% (373/688) for RT-PCR positive cases and 71.4% (679/950) for negative cases. Agreement was highest for RT-PCR negative cases labeled as *Normal (*50.4%, n= 479). Reader performance did not improve with clinical history or time between CXR and RT-PCR result.

**Conclusion:** At the time of presentation to the emergency department, emergency radiologist performance is non-specific for diagnosing COVID-19.

## Introduction

Despite lower sensitivity (69% versus 91%) of chest X Ray (CXR) compared to Chest CT and RT-PCR at the time of COVID-19 diagnosis (1), CXR is frequently used to screen for disease severity and patient triage pending the availability of final RT-PCR test results. For example, despite guidelines recommending use of CXR for monitoring rather than diagnosis (2,3), hospitals in Italy and Britain adopted CXR as a first line screening tool, (4,5), as it is cheap, portable, and eliminates the complexity of sanitizing CT rooms and infection prevention after patient scanning. (6) Initial studies early in the pandemic helped describe and understand the severity of findings of findings of COVID-19 on CXR (1,7–9). Wong et al evaluated radiographs from 69 patients and had 2 radiologists assign a severity score, as well as corresponding CT chest review (1). From these studies, we learned that peripheral consolidations were the most common CXR findings, peak radiographic findings occurred 10-12 days from symptom onset, and findings were predominantly in the bilateral lower lobes with a peripheral distribution(1,9).

Despite literature highlighting common CXR findings of COVID-19, diagnostic difficulty has increased as the COVID-19 pandemic evolved from sporadic outbreaks to widespread community transmission, altering the case mix of patients presenting to the emergency department (ED). This difficult is again renewed with the recent surge in cases due to the Delta variant. In this context, we examine the performance of emergency radiologists in diagnosing COVID-19 on CXR from the ED. Previous work by Murphy et al included 6 radiologists’ interpretation of 454 images, demonstrating specifies of 0.37 – 0.42 at the reader’s highest sensitivities, however the case distribution of this overseas dataset was not reported(10). In order to better delineate emergency radiologist performance on US data, we conducted a multi-institutional, multireader study in which ten emergency radiologists were presented with CXRs of patients presenting with respiratory or other thoracic symptoms, and evaluate their performance for COVID-19 diagnosis. Our study is representative of a commonly encountered workflow, wherein a person under investigation (PUI) may present to the ED with multiple symptoms, and the CXR is interpreted by an emergency radiologist or other non-thoracic radiologist.

## Materials and Methods

### Patients

Our institution’s Institutional Review Board reviewed and approved this retrospective study (STUDY00000506) and granted a waiver of HIPAA authorization and written informed consent. We extracted RT-PCR COVID results for all patients tested between March 2020 to June 2020 across 4 institutional hospitals. A list of chest radiographs obtained for these patients was extracted and then further filtered by Emergency Department location and date of radiograph within 3 days prior or 7 days after RT-PCR results (Figure 1). There were a mix of portable and two-view chest radiographs, therefore only AP and PA portable and non-portable views were used. The final sample size contained 984 images from patients who tested RT-PCR positive and 1,043 images from RT-PCR negative patients.

**Figure 1.**
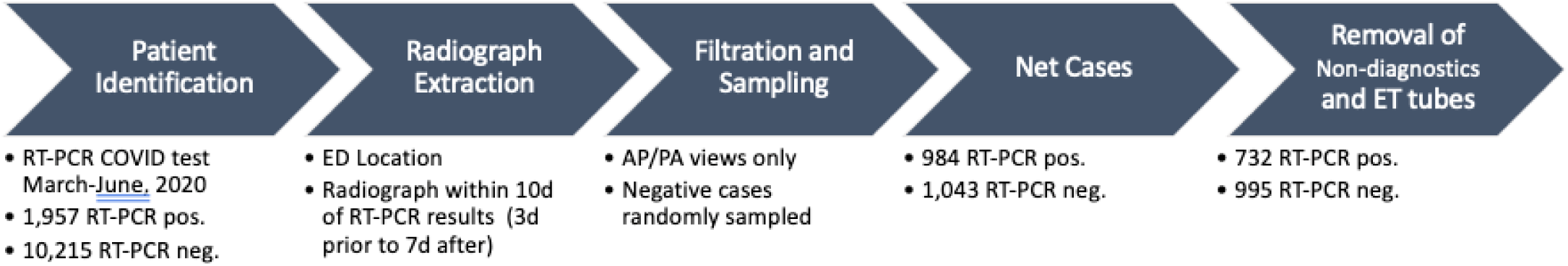
Workflow of patient identification for RT-PCR positive and RT-PCR negative radiographs, yielding 984 RT-PCR positive images and 1,043 RT-PCR negative images. Negative cases were much more prevalent and therefore were randomly sampled to yield a similar number to positive cases.

### Reader study

All de-identified images were uploaded to the online annotation platform MD.ai.(10) Ten radiologists with primary practice in the emergency department were recruited from four academic institutions and instructed to label images with the *single* best label from the following six choices: *Normal, COVID-19, Other - Infectious, Other - Noninfectious, Nondiagnostic Image*, or *Endotracheal Tube Present*. Nondiagnostic images were those that were either technically inadequate or of the wrong anatomic region. Readers were blinded to RT-PCR results and instructed to treat each image as a person under investigation (PUI) to reflect the common emergency department workflow where patient information may be unknown and symptoms such as cough, dyspnea, shortness of breath, or fever may overlap. 100 cases (50 RT-PCR positive and 50 RT-PCR negative) were randomly selected to be read by all 10 readers and the remaining 1927 cases were randomly assigned to be read by two readers. Therefore, reader interpreted 100 cases read by all readers and an addition 385-386 cases read by two readers, for a total of 485-486 cases per reader. Following annotation, all labels were downloaded in .*json* format and parsed.

For analysis, clinical history was extracted from the ‘History / Indication’ section of the radiology report and categorized as PUI, cough, infection, shortness of breath, chest pain, or other. The years of experience and fellowship training of each reader was also tracked.

### Statistical Analysis

Cases labeled as *non-diagnostic* were excluded, as were cases with *endotracheal tube* to decrease biased performance for the sickest patients. Descriptive statistical analysis was performed using the Pandas and scikit-learn Python library. (11) Data visualizations were generated using matplotlib and Seaborn in Python. (12,13) Interreader agreement was calculated using the Fleiss κ score as implemented in the *nltk*.*metrics*.*agreement* Python (14) module.

## Results

### Patient Characteristics

A total of 2027 radiographs were labeled, 984 RT-PCR positive (from 551 patients) and 1043 RT-PCR negative (from 1,043 patients). Table 1 summarizes the demographic distribution of our sample population. Mean age was similar between the groups at 58 ± 18 years for RT-PCR positive and 60 ± 19 for RT-PCR negative. 55.1% of RT-PCR positive patients and 50.1% of RT-PCR negative patients were between age 60-90 years. Genders distribution was nearly equal. The mean number of days between CXR and the RT-PCR test was 0 ± 1 day.

**Table 1:** Demographics distribution of patients included in the study. There was a relatively even split between male and female patients with a slight predominance of RT-PCR positive male patients. There was a majority of African American patients in both groups, however this was more pronounced for the RT-PCR positive group. Gender was unavailable for 2 cases and race and ethnicity was unavailable for 12 cases

|  | RT-PCR+ (n = 551) | RT-PCR- (n = 1043) |
| --- | --- | --- |
| **Gender and Age** |  |  |
| Male | 291 [52.8%] | 525 [50.4%] |
| Female | 258 [46.8%] | 517 [49.6%] |
| Unknown Gender | 2 [0.4%] | 1 [0.1%] |
| Age (years) | 58 ± 18 | 60 ± 19 |
| **Race** |  |  |
| African American | 370 [67.4%] | 533 [51.2%] |
| Caucasian or White | 114 [20.8%] | 437 [41.9%] |
| Asian | 11 [2.0%] | 35 [3.4%] |
| American Indian or Alaskan Native | 2 [0.4%] | 4 [0.4%] |
| Native Hawaiian or Other Pacific Islander | 1 [0.2%] | 1 [0.1%] |
| Multiple | - | 3 [0.3%] |
| Unavailable | 53 [9.6%] | 29 [2.8%] |
| **Ethnicity** |  |  |
| Non-Hispanic or Latino | 470 [85.6%] | 972 [93.3%] |
| Hispanic or Latino | 27 [4.9%] | 29 [2.8%] |
| Unreported, Unknown, Unavailable | 54 [9.8%] | 41 [3.8%] |

Patients were predominantly African American (n= 903, 56.6%), followed by White (n = 551, 34.6 %), and Asian (n = 46, 2.9%). There was a much higher prevalence of RT-PCR positive cases in African American patients (n= 370, 67.4%) compared to White patients (n = 114, 20.8%). In the RT-PCR negative group, 51.2 % (n= 533) of the patients were African American and 41.9 % (n=437) were white. Since we did not include the cases from the pediatric hospital, we had very few patients under 20 and no patients under 10 years of age.

14 RT-PCR positive radiographs and 40 RT-PCR negative radiographs were excluded due to the *non-diagnostic* label, with reasons including incompletely visualized thoracic cavity, wrong anatomic region (i.e. mislabeled metadata), or excessive over/under-penetration of the radiograph. An additional 244 radiographs in the RT-PCR positive group and 7 patients in the RT-PCR negative group were excluded due to presence of endotracheal tube. This yielded 1727 (732 RT-PCR positive and 995 RT-PCR negative) remaining radiographs for statistical analysis. 89 of these had been read by all 10 readers and 1,638 were read by two readers.

### Emergency Radiologist Characteristics

Ten radiologists who work in the ED at 4 academic institutions performed the reader study, with 5 readers from institution 1, 3 readers from institution 2, 1 reader from institution 3, and 1 reader from institution 4. Years of experience after completion of training ranged from 1-13 years. Two readers were trained in breast imaging, two in cardiothoracic imaging, two in body imaging, one in neuroradiology, one in emergency radiology, and two had no fellowship training (Table 2).

**Table 2:** Summary of radiologist readers. All readers practiced in the Emergency Division and had varied years of experience and fellowship training. Specificity was high for all users whereas sensitivity varied widely from 0.12 to 0.54 with a trend for decreased specificity as sensitivity increased. Sn = Sensitivity; Sp = Specificity; PPV = positive predictive value; NPV = negative predictive value

| Reader | Institution | Fellowship | Years in Practice | Sn | Sp | PPV | NPV |
| --- | --- | --- | --- | --- | --- | --- | --- |
| 1 | Institution 1 | Body | 7 | 0.273 | 0.983 | 0.942 | 0.577 |
| 2 | Institution 2 | Emergency | 11 | 0.247 | 0.991 | 0.968 | 0.559 |
| 3 | Institution 1 | Breast | 3 | 0.074 | 0.996 | 0.947 | 0.507 |
| 4 | Institution 1 | Cardiothoracic | 5 | 0.335 | 0.984 | 0.951 | 0.610 |
| 5 | Institution 3 | Breast | 3 | 0.387 | 0.974 | 0.941 | 0.598 |
| 6 | Institution 1 | Body | 10 | 0.374 | 0.984 | 0.955 | 0.635 |
| 7 | Institution 2 | None | 13 | 0.416 | 0.972 | 0.932 | 0.643 |
| 8 | Institution 4 | Cardiothoracic | 1 | 0.260 | 0.980 | 0.922 | 0.597 |
| 9 | Institution 2 | None | 11 | 0.542 | 0.915 | 0.854 | 0.685 |
| 10 | Institution 1 | Neuro | 3 | 0.118 | 0.996 | 0.967 | 0.519 |

### Reader study

Overall agreement between the 10 readers on 89 cases was fair, with a Fleiss score of 0.36 for RT-PCR positive cases and 0.46 for RT-PCR negative cases (Figure 2). Agreement was slightly higher for radiologists with >5 years experience (0.38) as compared to radiologists with <= 5 years experience (0.33), but this trend was not observed for RT-PCR negative cases. Overall sensitivity was 0.303, specify of 0.978, PPV of 0.938, and NPV of 0.593. We did not note any trends in performance according to fellowship training, and the thoracic-trained radiologist did not perform significantly better than the others. The two readers with the highest sensitivity had the greatest years of experience (13 and 11 years), but specificity suffered slightly (Table 2).

**Figure 2.**
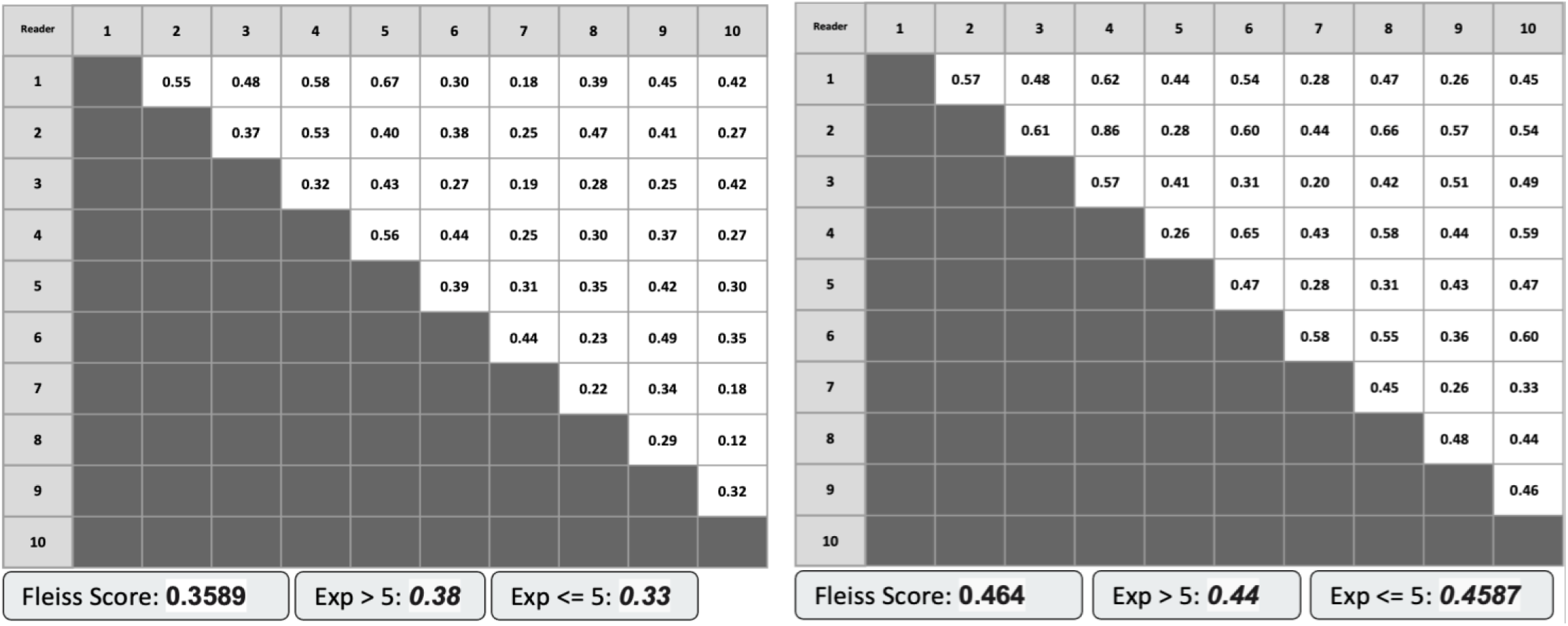
Interrater agreement of the 10 radiologists for 44 RT-PCR positive cases (top) and 45 RT-PCR negative cases (bottom). Overall agreement was higher for negative cases. For positive cases, agreement was slightly higher for radiologists with >5 years experience than those with <=5 years experience. This trend was not observed for RT-PCR negative cases.

Agreement of the 89 RT-PCR positive and RT-PCR negative cases annotated by all 10 readers are shown as swarm plots (Figure 3) whose shapes represent the distribution of reader agreements. Results for the RT-PCR positive patients show a wide distribution for all labels, with most readers agreeing that the CXRs were not *Normal*, but relatively little agreement *COVID-19, Other - Infectious* and *Other - Noninfectious* labels. RT-PCR negative cases demonstrate an overall tighter distribution for all labels indicating that readers have a high level of agreement (>5 readers agreed) for RT-PCR negative cases. Agreement was high for cases labeled *Normal* and in *not* labeling cases as *COVID-19* and *Other - Infectious*.

**Figure 3.**
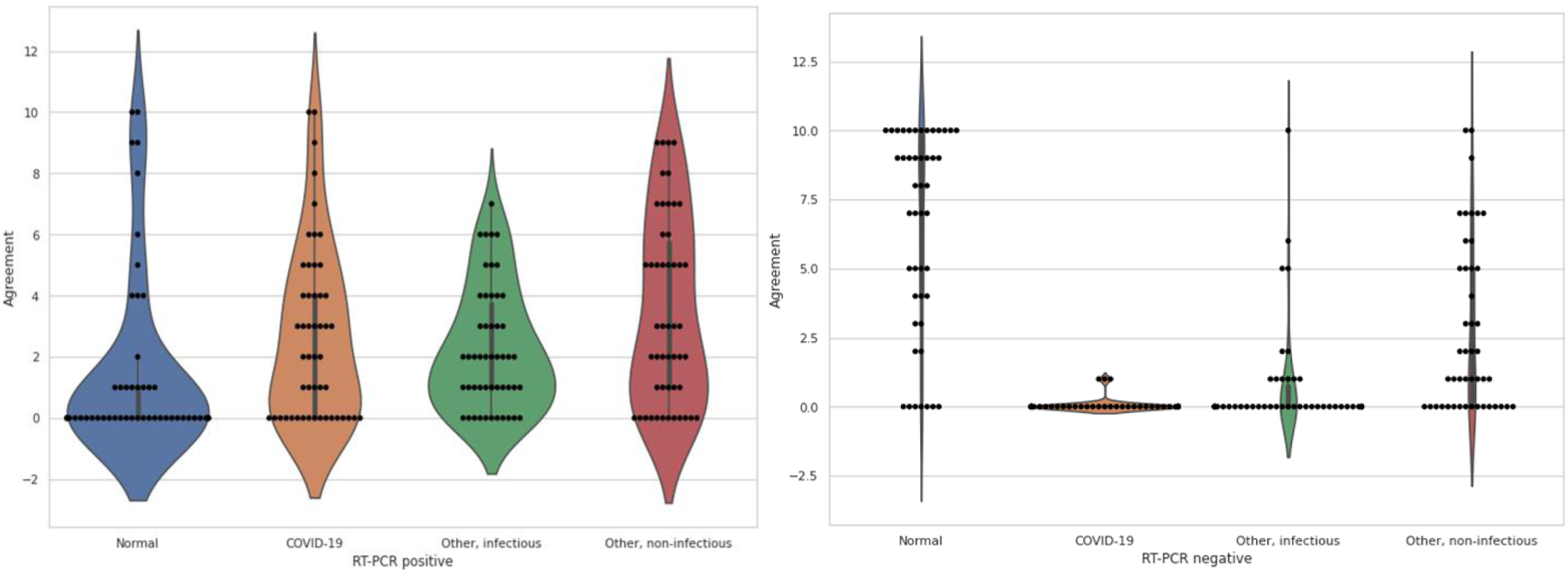
*Left*: Swarm plot showing the agreement between 10 radiologists on 44 RT-PCR positive chest radiographs. Radiologists were asked to assign one of four labels per radiograph, denoted along the x-axis. The y-axis denotes the number of radiologists that assigned a given label to the case, and each black dot represents one case with the specified level of agreement. A dot on the Y-axis at 10 indicates all 10 radiologists assigned the label and a dot at zero indicates no radiologists assigned the label. Overall agreement was poor for most cases with readers agreeing only that cases were not *Normal. Right*: Agreement between 10 radiologists on 45 RT-PCR negative chest radiographs. There was overall good agreement for *Normal* cases and for labeling cases as not *COVID-19* or *Other - Infectious*.

The distribution for 1638 cases labeled by two readers is shown as a confusion matrix in Figure 4. We observed a similar pattern of agreement for RT-PCR negative cases, with 50.4% (479/950) labeled as Normal by both readers and only 0.6% (6/950) labeled as *COVID-19* by both readers. For RT-PCR positive cases, only 15.9% (110/688) cases were labeled as *COVID-19* by both readers and 17.8% (123/688) of cases were judged as normal by both readers suggesting that the patient likely did not yet have radiographic evidence of disease. Overall agreement between both readers was 54.2% (373/688) for RT-PCR positive cases and 71.4% (679/950) for RT-PCR negative cases. Sample images for RT-PCR positive and negative cases in which both readers agreed and disagreed are shown in Figure 5.

**Figure 4.**
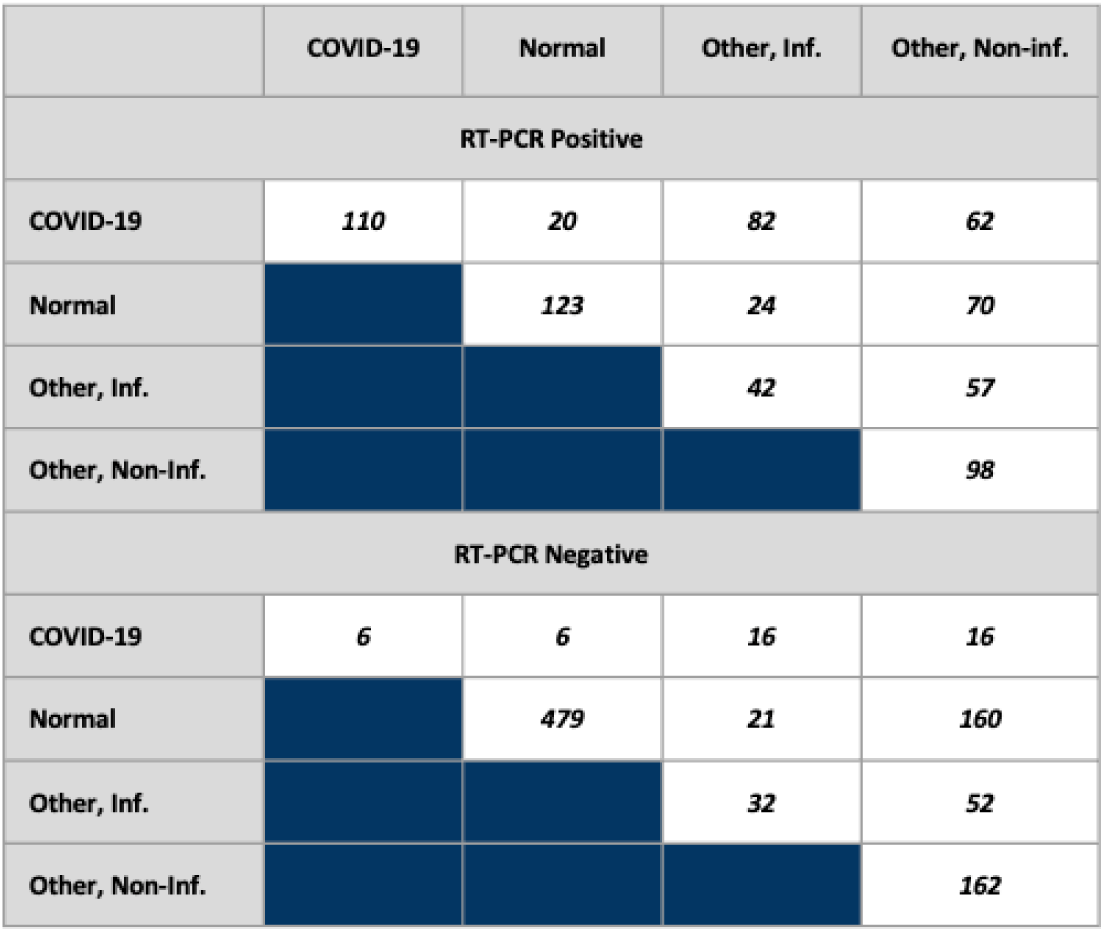
Distribution of labels for 688 RT-PCR positive and 950 RT-PCR negative cases, demonstrating highest agreement for *normal* in RT-PCR - cases, and poor agreement for *COVID-19* in RT-PCR + cases.

**Figure 5.**
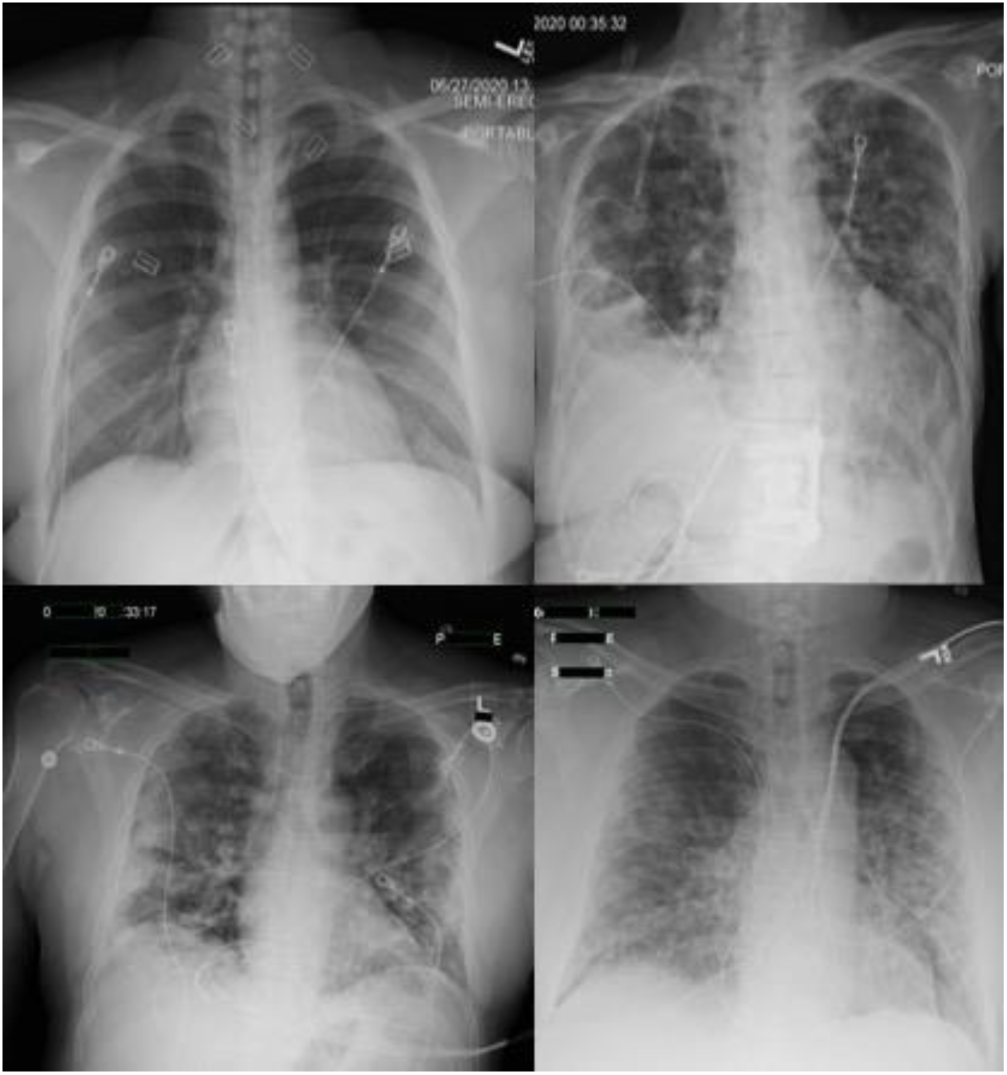
(A) RT-PCR negative case in which both readers agreed the case was *normal*. (B) RT-PCR negative case in which readers disagreed, with one reader labeling *COVID-19* and the other labeling *Other - Infectious*. (C) RT-PCR positive case in which both readers agreed on *COVID-19*. (D) RT-PCR positive case in which readers disagreed, with one reader labeling *COVID-19* and the other labeling *Other – Noninfectious*.

RT-PCR positive patients with multiple exams were selected for sub-analysis, yielding 136 patients with between two and five exams (patients with >5 exams were excluded). Assigned labels from two readers are plotted as a function of days between RT-PCR test and CXR (Figure 6). Agreement between labels remained poor, and there was no trend in interreader agreement with respect to time for in these cases.

**Figure 6.**
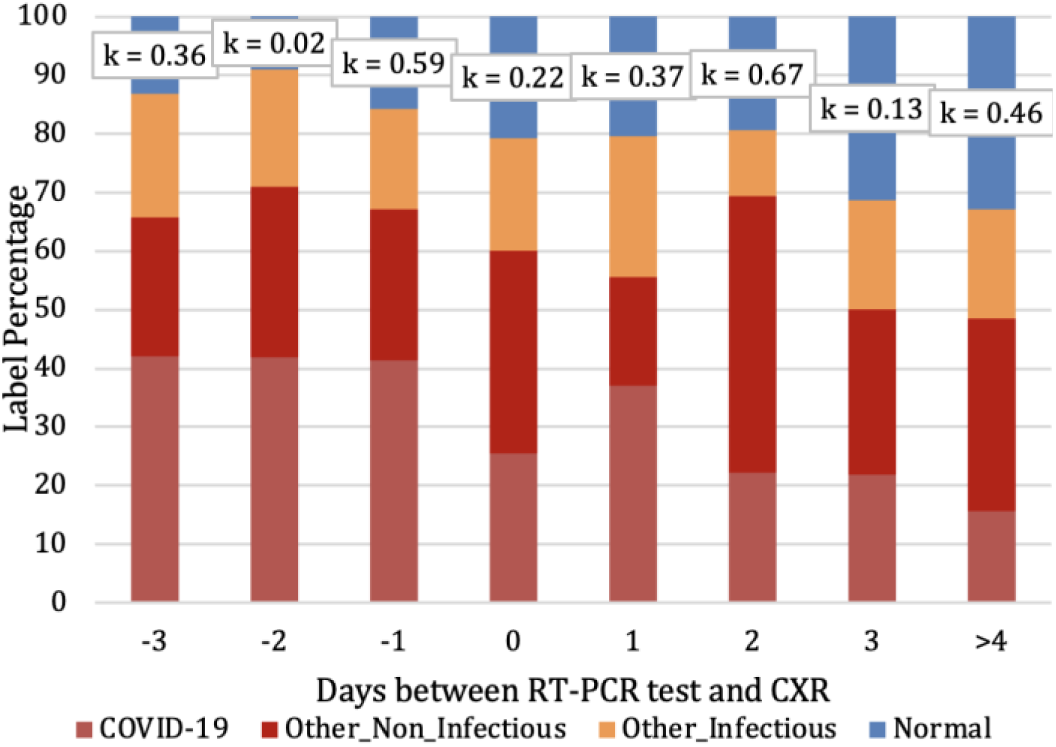
Distribution of labels for RT-PCR positive cases in 136 patients with between two and five exams. Kappa score (k) is listed at the top of each bar. Agreement remains poor regardless of days between RT-PCR testing and CXR, specifically there is no improvement in reader performance for CXR that occurred several days after RT-PCR testing.

Distribution of clinical indications were relatively even between RT-PCR positive and negative cases, with shortness of breath and person under investigation (PUI) being most common (Figure 7). Chest pain was more frequent in RT-PCR negative patients (22.6%, 214/950) than RT-PCR positive (7.0%, 48/688). Clinical indication did not significantly impact the assigned label for either RT-PCR positive or negative cases, regardless of whether the readers agreed with one another. We did observe that for the clinical indication of *cough* for both RT-PCR positive and negative patients, both readers agreed on *Normal* more often than other labels. We also observed that for cases in which radiologists disagreed, the *Other - Non-infectious* label was frequently selected.

**Figure 7.**
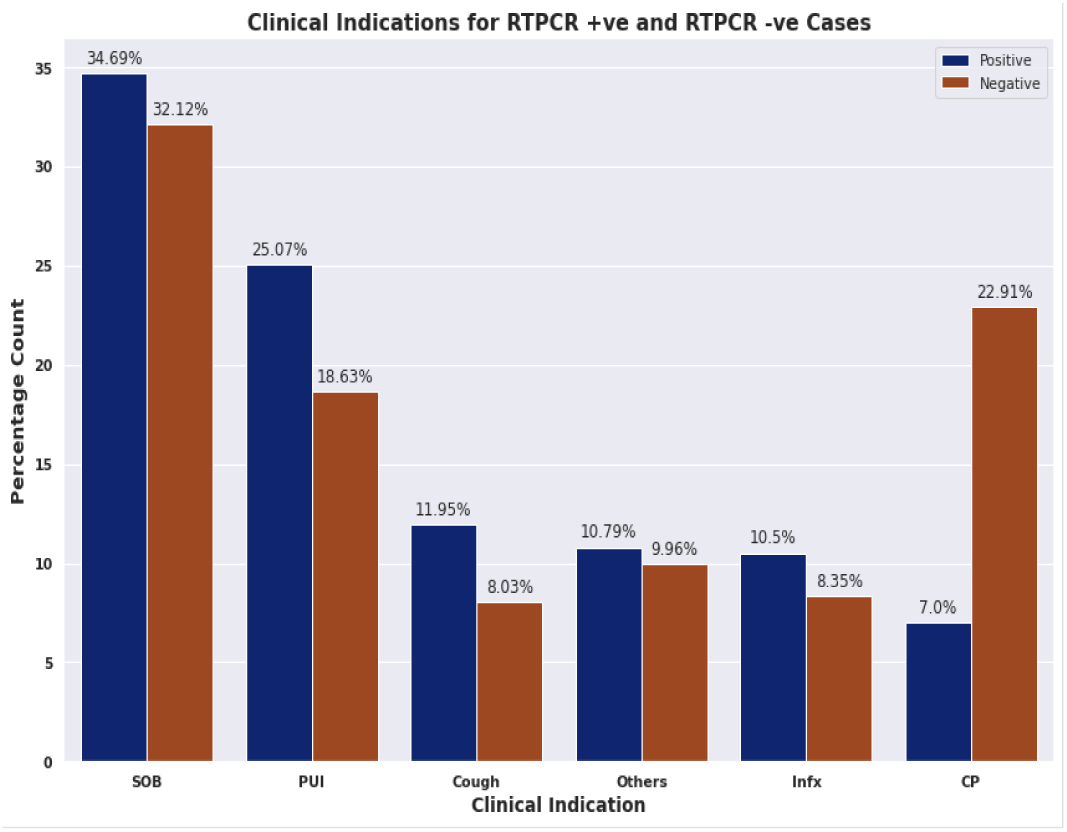
Distribution of clinical indications as extracted from the ‘Clinical History’ section of reports for RT-PCR positive and negative cases. Symptom severity was not available. Shortness of breath and persons under investigation were most common in both groups. Chest pain was more common in the RT-PCR negative cases. SOB = shortness of breath; PUI = persons under investigation; Infx = infection; CP = chest pain.

## Discussion

There were many challenges to COVID-19 diagnosis early in the pandemic due to lack of readily available and rapid RT-PCR testing, which led to heavy reliance on imaging for diagnosis. This limitation may still exist in underserved regions both domestically and internationally or as new waves such as the recent surge due to the Delta variant occur and strain healthcare resources. Our study demonstrates three major findings, 1) that there is low utility of CXRs in diagnosing patients who will be COVID-19 positive, 2) clinical history is not useful in improving the radiologist performance for COVID-19 diagnosis, and 3) CXRs are more useful for excluding COVID-19 diagnosis with a consistent level of performance across a diverse group of radiologists. There was a very low rate of assigning COVID-19 labels for RT-PCR negative patients, totaling 0.6% (6/950) when two radiologists agreed and 4.0% (38/950) when they disagreed. This observation persists even with the introduction of clinical history of chest pain, cough, infection, PUI or shortness of breath.

For RT-PCR positive patients, we find that radiologists have poor interrater agreement of labels (Fleiss score 0.36), and nonspecific performance for diagnosing COVID-19 with wide distribution of the remaining labels and low sensitivity for detection. Two readers agreed on a COVID-19 diagnosis only 15.9% (110/688) of the time, and one of two readers labeled COVID-19 23.8% (164/688) of the time.

This performance does not improve even when the clinical history is provided, and improves only slightly with >5 years of experience among the readers. We also did not note any increase in COVID-19 diagnostic performance with respect to time elapsed between a positive RT-PCR result and CXR exam. This was surprising as severity of COVID-19 radiographic findings would be expected to increase with time. However, this lack of correlation could be attributed to the fact that patients are symptomatic for varying amounts of time and with varying severity before presentation and testing, owing to the heterogenous effects of COVID-19 on the general population.

Overall, our results are consistent with the known literature on the diagnostic capabilities of CXR for the diagnosis of COVID-19 pneumonia(1,2). They also reinforce with multiple expert panel recommendations which recommend against routine use of CXR as a COVID-19 screening test, but rather as a tool to monitor severity of disease (3).

Our study has several limitations. In our study design, we focused only on CXRs from the ED, and hence excluded the longitudinal follow-up of imaging that may occur when patients are admitted in the hospital. These later studies would likely have more radiographic abnormalities and increased accuracy of COVID-19 detection, but RT-PCR results are more likely to be available after admission and diagnosis of COVID-19 via CXR becomes less important. It is also possible that the long incubation time for COVID-19 (up to 10 days) means that RT-PCR positive patients could be asymptomatic and therefore a normal CXR is expected in many patients. We were unable to classify patients by disease severity using available data, and it is possible that stratifying radiologists performance by disease severity could demosntrate improved performance for more severe cases. Radiologist performance could also potentially be improved with specific training in COVID-19 diagnosis on radiography. Lastly, this study did not include chest CT findings, which could help resolve some of the findings from *Other – Noninfectious* or *Other - Infectious* to *COVID-19*. However, in the typical ED workflow, CXR is performed before chest CT, and hence the additional imaging for clarification would not be available at the time of CXR interpretation.

In conclusion, our study mirrored to the ambulatory patient journey can provide insight into the design of patient triage for COVID-19 in the ED. Such triage workflows are more important especially in a pandemic where resources are limited, and ED patient flow is important. Our sample size is filtered to the presenting radiograph at the time of diagnosis, and clinical history is provided to the readers who work in the ED. We conclude that emergency radiologists have moderate performance for excluding COVID negative patients, but when initial results are abnormal then an RT-PCR result is necessary for the final patient diagnosis. The clinical history did not improve the performance of the radiologists. We also did not specifically study the performance of thoracic-trained radiologists. As image-based algorithm development increases for COVID-19 diagnosis, we urge caution in interpreting the results of these algorithms and their comparison to radiologist performance. We plan to release the annotated CXR dataset to aid in such evaluation.

## Data Availability

The data produced is available upon reasonable request to the authors.

## Acknowledgments

The authors would like to acknowledge Dr. Elizabeth Krupinski for her assistance with study design and analysis recommendation for the reader analysis.

